# Prevalence of suicidal ideation and its associated factors among school-going adolescents in northern Bangladesh: a cross-sectional study

**DOI:** 10.64898/2026.10.07.26365009

**Authors:** Abdul Mannan, Mohammad Delwer Hossain Hawlader, S M Saidur Rahman Mashreky

**Author notes:** Corresponding author: Md. Abdul Mannan.

## Abstract

Adolescent suicidal ideation is a critical public health issue, yet empirical evidence on its prevalence and associated factors among adolescents in northern Bangladesh remains limited. This study estimated the 12-month prevalence of suicidal ideation and examined its associated factors among school-going adolescents in Nilphamari District, Bangladesh. A cross-sectional survey was conducted between May and June 2026 among 365 adolescents aged 13–19 years across eight secondary schools and colleges. Self-administered questionnaires assessed suicidal ideation alongside validated tools for depression (PHQ-9), anxiety (GAD-7), and perceived social support (MSPSS). Factors associated with suicidal ideation were examined using multivariable logistic regression. The 12-month prevalence of suicidal ideation was 30.7% (112/365; 95% CI 26.0%–35.7%). In the primary multivariable model, moderate depression (AOR = 6.05, 95% CI 1.03-35.38; *p* = 0.046) and moderately severe depression (AOR = 6.86, 95% CI 1.05-44.62; *p* = 0.044) were associated with higher odds of suicidal ideation compared to minimal depression. However, this association was not statistically significant in a sensitivity analysis using the PHQ-8 score after excluding Item 9 (AOR = 1.06, 95% CI: 0.96-1.17; p = 0.228). Loneliness “often” showed the strongest association with suicidal ideation (AOR = 10.59, 95% CI: 2.53-44.35; p = 0.001). Suicidal ideation was also associated with truancy of 6–9 days (AOR 4.44, 95% CI 1.24–15.90; p = 0.022), truancy of ≥10 days (AOR 3.77, 95% CI 1.34–10.59; p = 0.012), intentional self-harm (AOR = 2.51, 95% CI: 1.17–5.41; p = 0.018), and physical harm exposure (AOR = 2.63, 95% CI: 1.24–5.57; p = 0.012). Maternal secondary education was associated with lower odds of suicidal ideation (AOR = 0.34, 95% CI: 0.12–0.99; p = 0.048). Suicidal ideation was common among school-going adolescents in northern Bangladesh, affecting nearly one in three participants. Early school-based identification and support for adolescents experiencing mental-health and psychosocial difficulties, together with family engagement and strengthened suicide-prevention services, are warranted.

## Introduction

Adolescent mental health is a major global public health priority, with suicidal ideation serving as a primary indicator of severe psychological distress during this critical developmental stage. As suicide remains one of the leading causes of death among young people worldwide, early identification of suicidal thoughts is crucial [1–3]. Although suicidal ideation does not consistently progress to suicidal behaviour, it reliably signals underlying depression, anxiety, and exposure to adverse social environments [4,5]. This burden falls disproportionately on resource-limited settings; nearly nine out of ten global adolescent suicides occur in low- and middle-income countries (LMICs), where nearly 90% of the world’s adolescents reside [1]. In these contexts, adolescent vulnerability is further exacerbated by limited access to mental health services, socioeconomic hardship, exposure to violence and pervasive stigma surrounding mental health and help-seeking behaviours [6,7].

While epidemiological data on adolescent suicidal ideation in Bangladesh is growing, substantial gaps remain regarding its local magnitude and associated factors. A recent systematic review reported a pooled 12-month prevalence of suicidal thoughts of 15.6% among Bangladeshi students, though estimates varied considerably across populations [8]. Individual studies reflect this divergence, with prevalence estimates ranging from 11.7% in a national school-based survey [9] and 3.2% in Sirajganj District [10] to 19.6% among rural adolescents in Narail district [11].

Suicidal ideation among adolescents is multifactorial, driven by an interplay of psychological, behavioural, interpersonal, and socioeconomic factors. While depression and anxiety are well-established psychological correlates, factors such as family conflict, academic pressure, self-harm, bullying, violence, and loneliness further heighten vulnerability [4,5,9,12,13]. Conversely, strong perceived social support and positive parental characteristics such as higher maternal education have been associated with more favourable adolescent mental-health outcomes, including through emotional communication and help-seeking. [14–16].

Despite these recognised correlates, empirical evidence on adolescent suicidal ideation in northern Bangladesh remains sparse. Specifically, little is known regarding the combined influence of clinical, interpersonal, and family-level factors in this socioeconomically distinct region. Nilphamari District offers a critical setting to examine these dynamics among secondary school and college students. Generating localised data from this population is essential for identifying high-risk adolescents and designing contextually tailored, school- and community-based suicide prevention strategies.

Therefore, this study aimed to estimate the prevalence of suicidal ideation and identify its associated factors among school-going adolescents in Nilphamari District, northern Bangladesh.

## Methods

### Study design and setting

A school-based cross-sectional study was conducted among school-going adolescents in Nilphamari District, Bangladesh. Two upazilas (Nilphamari Sadar and Dimla) were randomly selected from six upazilas. Eight co-educational secondary schools and colleges were selected randomly. Data were collected between May 12 and June 11, 2026.

### Participants and sampling

The study population comprised school-going adolescents aged 13–19 years enrolled in Grades 8-12. Participants were eligible if they were present during data collection, able to complete the self-administered questionnaire independently, and provided written informed assent.

A multistage sampling approach was employed. Assuming a 15% anticipated prevalence of suicidal ideation, a 95% confidence level, a 5% margin of error, a design effect of 1.5, and a 10% non-response allowance, the target sample size was calculated as 324. A total of 371 adolescents participated; six questionnaires were excluded because of incomplete responses, leaving 365 participants for the final analysis.

### Variables and measurements

**Primary outcome:** 12-month suicidal ideation was assessed using a single-item adapted from the Global School-based Student Health Survey (GSHS): *“During the past 12 months, did you ever seriously consider attempting suicide?”* (Responses were recorded as binary: Yes/No).

**Explanatory variables:** Data were collected on socio-demographic, socioeconomic, socio-environmental, psychosocial, behavioural, and health-related factors. Depression severity was evaluated using the Patient Health Questionnaire-9 (PHQ-9; minimal: 0–4, mild: 5–9, moderate: 10–14, moderately severe: 15–19, severe: 20–27). Anxiety severity was measured using the Generalised Anxiety Disorder-7 (GAD-7; minimal: 0–4, mild: 5–9, moderate: 10–14, severe: 15–21). Perceived social support was assessed using the Multidimensional Scale of Perceived Social Support (MSPSS) categorised as low, moderate, or high support.

### Data collection and quality assurance

Data were collected using a structured, anonymous, paper-based, self-administered questionnaire supervised by the principal investigator. The questionnaire was pre-tested among 15 adolescents in a non-study institution to refine clarity and sequencing. Completed questionnaires were verified immediately for completeness, entered into Microsoft Excel using predefined coding, and audited prior to statistical analysis.

### Statistical analysis

Data were analysed using Stata Statistical Software Release 17 (StataCorp LLC, College Station, TX, USA). Categorical variables were summarised using frequencies and percentages, while continuous variables as means with standard deviations (SD).

Pearson’s chi-square test was used to assess bivariate associations between explanatory variables and suicidal ideation, and binary logistic regression to estimate crude odds ratios (CORs) with 95% confidence intervals (CIs).

Multivariable binary logistic regression was performed to examine factors associated with suicidal ideation, with adjusted odds ratios (AORs) and 95% CIs reported. To optimise model stability, sparse categories for parental homework checking and knowledge of free-time activities were collapsed into “Rarely or Never”. Multicollinearity was evaluated using Variance Inflation Factors (VIF). Model goodness-of-fit was evaluated via the Hosmer–Lemeshow test, and discrimination was assessed using receiver operating characteristic (ROC) curves and area under the curve (AUC). Statistical significance was set at two-sided p < 0.05.

A sensitivity analysis was conducted using the PHQ-8 score after excluding PHQ-9 Item 9, which addresses thoughts of death or self-harm and may conceptually overlap with the suicidal ideation outcome. The PHQ-8 score was entered as a continuous variable in place of depression severity, with all other covariates retained as in the primary model. This study is reported according to the Strengthening the Reporting of Observational Studies in Epidemiology (STROBE) guideline (S1 Checklist).

### Ethics statement

Ethical approval was obtained from the Institutional Review Board (IRB) of North South University (Ref: 2026/OR-NSU/IRB/0511). Administrative permissions were granted by regional and district education authorities and participating institution heads. Institutional permission and participant written informed assent were utilised in lieu of parental consent for this minimal-risk, school-based study, as parental consent was waived by the IRB. Anonymity and voluntary participation were preserved throughout.

## Results

### Characteristics of the study participants

A total of 365 school-going adolescents aged 13–19 years (mean age: 16.55 ± 1.71 years) were included in the final analysis. Females comprised 52.9% (n = 193) of the sample. Participants were evenly distributed across institution types (51.8% school, 48.2% college) and institution locations (51.0% urban, 49.0% rural), though most resided in rural areas (81.6%).

Regarding mental health, mean scores for depression (PHQ-9), anxiety (GAD-7), and perceived social support (MSPSS) were 9.92 ± 5.04, 8.43 ± 5.13, and 54.78 ± 13.14 respectively. Over half of the participants exhibited moderate to severe depression (50.7%) and close to four in ten showed moderate to severe anxiety (38.9%). Regarding behavioural and health risk factors, 23.6% reported a history of intentional self-harm, 23.6% experienced bullying, 26.0% experienced physical harm in the past 12 months, and 26.8% reported frequent loneliness. Detailed sample characteristics are presented in Table 1.

**Table 1.** Socio-demographic, mental health, psychosocial and behavioural characteristics of school-going adolescents in Nilphamari District, Bangladesh (N = 365)

| Characteristic | Category | n (%) / Mean ± SD |
| --- | --- | --- |
| Age in years | Mean ± SD (Range) | 16.55 ± 1.71 (13–19) |
| Sex | Male | 172 (47.1) |
|  | Female | 193 (52.9) |
| Class level | Grade 8 | 55 (15.1) |
|  | Grade 9 | 71 (19.5) |
|  | Grade 10 | 63 (17.3) |
|  | Grade 11 | 87 (23.8) |
|  | Grade 12 | 89 (24.4) |
| Institution type | School | 189 (51.8) |
|  | College | 176 (48.2) |
| Institution location | Urban | 186 (51.0) |
|  | Rural | 179 (49.0) |
| Residence area | Urban | 67 (18.4) |
|  | Rural | 298 (81.6) |
| Family size | $\leq 3$ | 26 (7.1) |
|  | 4–5 | 249 (68.2) |
| | $\geq 6$ | 90 (24.7) |
| Perceived family economic status | Low | 62 (17.0) |
|  | Middle | 289 (79.2) |
|  | High | 14 (3.8) |
| Mother's education | No Education | 38 (10.4) |
|  | Primary | 116 (31.8) |
|  | Secondary | 150 (41.1) |
|  | Higher Secondary | 45 (12.3) |
|  | >Higher Secondary | 16 (4.4) |
| Depression severity | Minimal | 58 (15.9) |
|  | Mild | 122 (33.4) |
|  | Moderate | 118 (32.3) |
|  | Moderately Severe | 48 (13.2) |
|  | Severe | 19 (5.2) |
| PHQ-9 total score | Mean ± SD | 9.92 ± 5.04 |
| Anxiety severity | Minimal | 99 (27.1) |
|  | Mild | 124 (34.0) |
|  | Moderate | 92 (25.2) |
|  | Severe | 50 (13.7) |
| GAD-7 total score | Mean ± SD | 8.43 ± 5.13 |
| Perceived social support | Low | 29 (8.0) |
|  | Moderate | 205 (56.2) |
|  | High | 131 (35.9) |
| MSPSS total score | Mean ± SD | 54.78 ± 13.14 |
| Parent-child relationship | Very Good | 106 (29.0) |
|  | Good | 140 (38.4) |
|  | Average | 108 (29.6) |
|  | Poor | 4 (1.1) |
|  | Very Poor | 7 (1.9) |
| Truancy (past 30 days) | 0 Days | 163 (44.7) |
|  | 1–2 Days | 89 (24.4) |
|  | 3–5 Days | 44 (12.1) |
|  | 6–9 Days | 26 (7.1) |
| | $\geq 10$ Days | 43 (11.8) |
| Academic pressure | Low | 44 (12.1) |
|  | Moderate | 111 (30.4) |
|  | High | 128 (35.1) |
|  | Very High | 82 (22.5) |
| Parental homework/school activity checking | Always | 89 (24.4) |
|  | Most of the Time | 104 (28.5) |
|  | Sometimes | 135 (37.0) |
|  | Rarely or Never | 37 (10.1) |
| Parental knowledge of free-time activities | Always | 96 (26.3) |
|  | Most of the Time | 109 (29.9) |
|  | Sometimes | 85 (23.3) |
|  | Rarely or Never | 75 (20.6) |
| Conflict with parents/guardians | Never | 129 (35.3) |
|  | Rarely | 136 (37.3) |
|  | Sometimes | 72 (19.7) |
|  | Often | 28 (7.7) |
| Family shouting during disagreements | Never | 80 (21.9) |
|  | Rarely | 160 (43.8) |
|  | Sometimes | 85 (23.3) |
|  | Often | 40 (11.0) |
| Family history of suicide | Yes | 61 (16.7) |
|  | No | 210 (57.5) |
|  | Don't Know | 94 (25.8) |
| Intentional self-harm | No | 279 (76.4) |
|  | Yes | 86 (23.6) |
| Bullying experience (past 12 months) | No | 279 (76.4) |
|  | Yes | 86 (23.6) |
| Physical harm (past 12 months) | No | 270 (74.0) |
|  | Yes | 95 (26.0) |
| Loneliness (past 30 days) | Never | 58 (15.9) |
|  | Rarely | 117 (32.1) |
|  | Sometimes | 92 (25.2) |
|  | Often | 64 (17.5) |
|  | Always | 34 (9.3) |
| Chronic illness | No | 325 (89.0) |
|  | Yes | 40 (11.0) |

### Prevalence of suicidal ideation

The overall 12-month prevalence of suicidal ideation was 30.7% (112/365; 95% CI: 26.0%–35.7%) (Fig 1).

**Fig 1.**
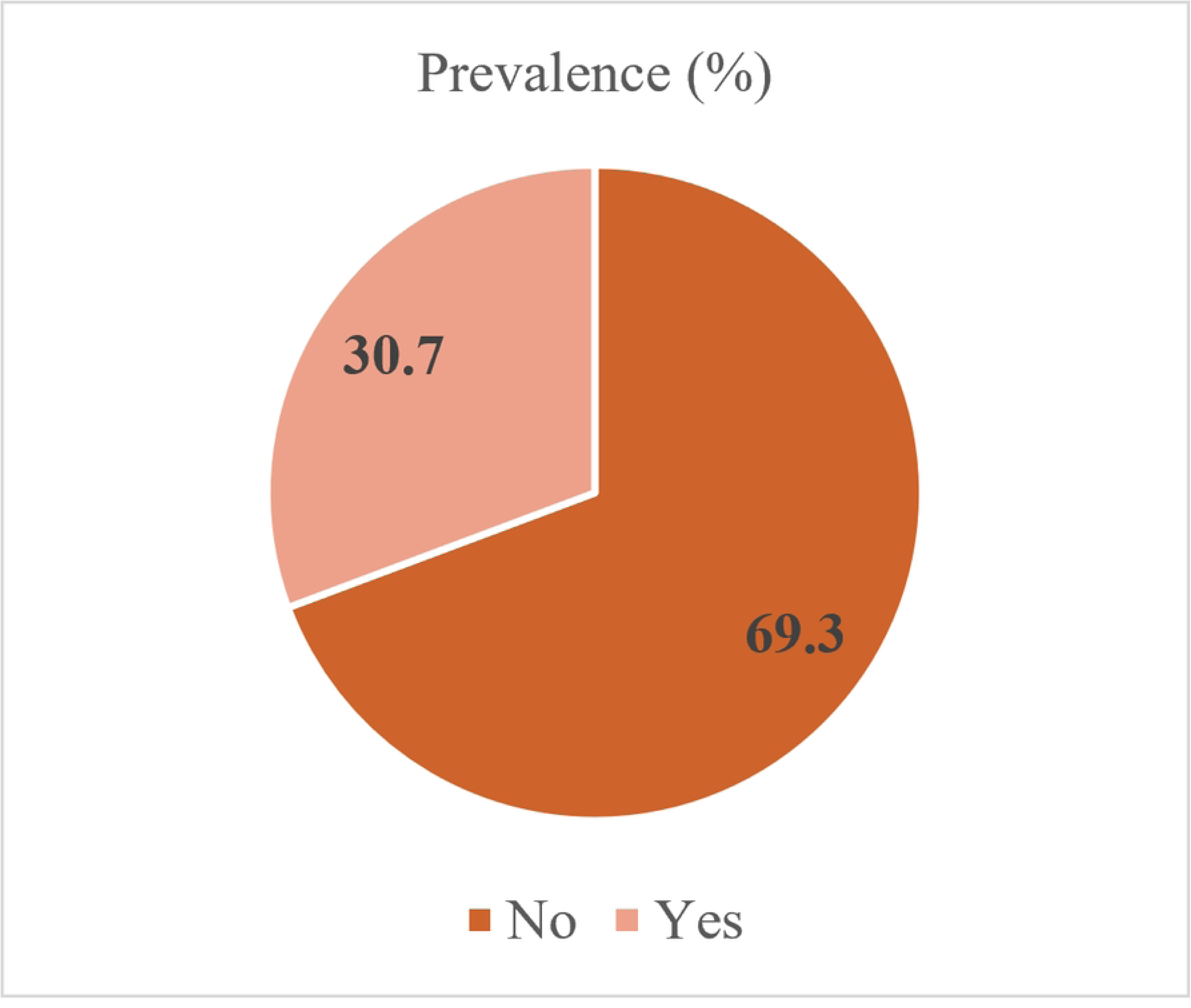
Prevalence of Suicidal Ideation among the Study Participants (N = 365)

### Bivariate associations with suicidal ideation

Bivariate chi-square analyses indicated that suicidal ideation was significantly more prevalent among adolescents living in urban areas (43.3% vs. 27.9%, p = 0.013), those from smaller families (≤ 3 members: 53.8%, p = 0.028), and those whose mothers had no formal education or higher education (p = 0.021). The prevalence of suicidal ideation increased progressively with depression severity, from 3.4% among adolescents with minimal depression to 78.9% (p < 0.001) among those with severe depression, and with anxiety severity, from 10.1% to 70.0% (p < 0.001) respectively. Significant associations were also observed for parent-child relationship, truancy, academic pressure, parental monitoring (homework check/knowledge of free time activities) conflict with parents or guardians, family shouting during disagreements, family history of suicide, intentional self-harm, bullying experience, physical harm, loneliness, and chronic illness (all p < 0.05; Table 2). Perceived social support, sex, grade level, institution type, and institution location were not significantly associated with suicidal ideation in bivariate testing. Additional bivariate findings are presented in S1 Table.

**Table 2.**
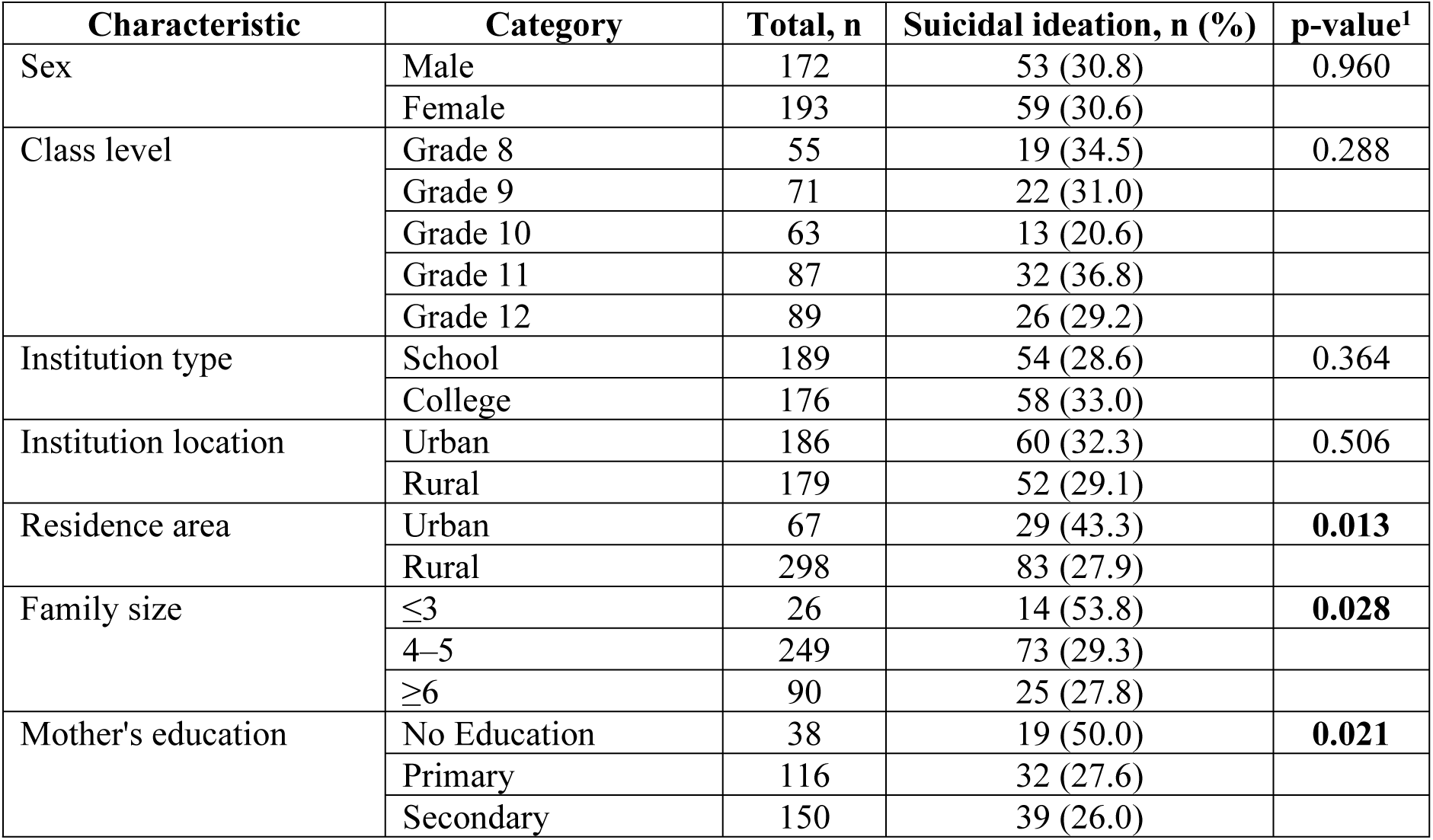
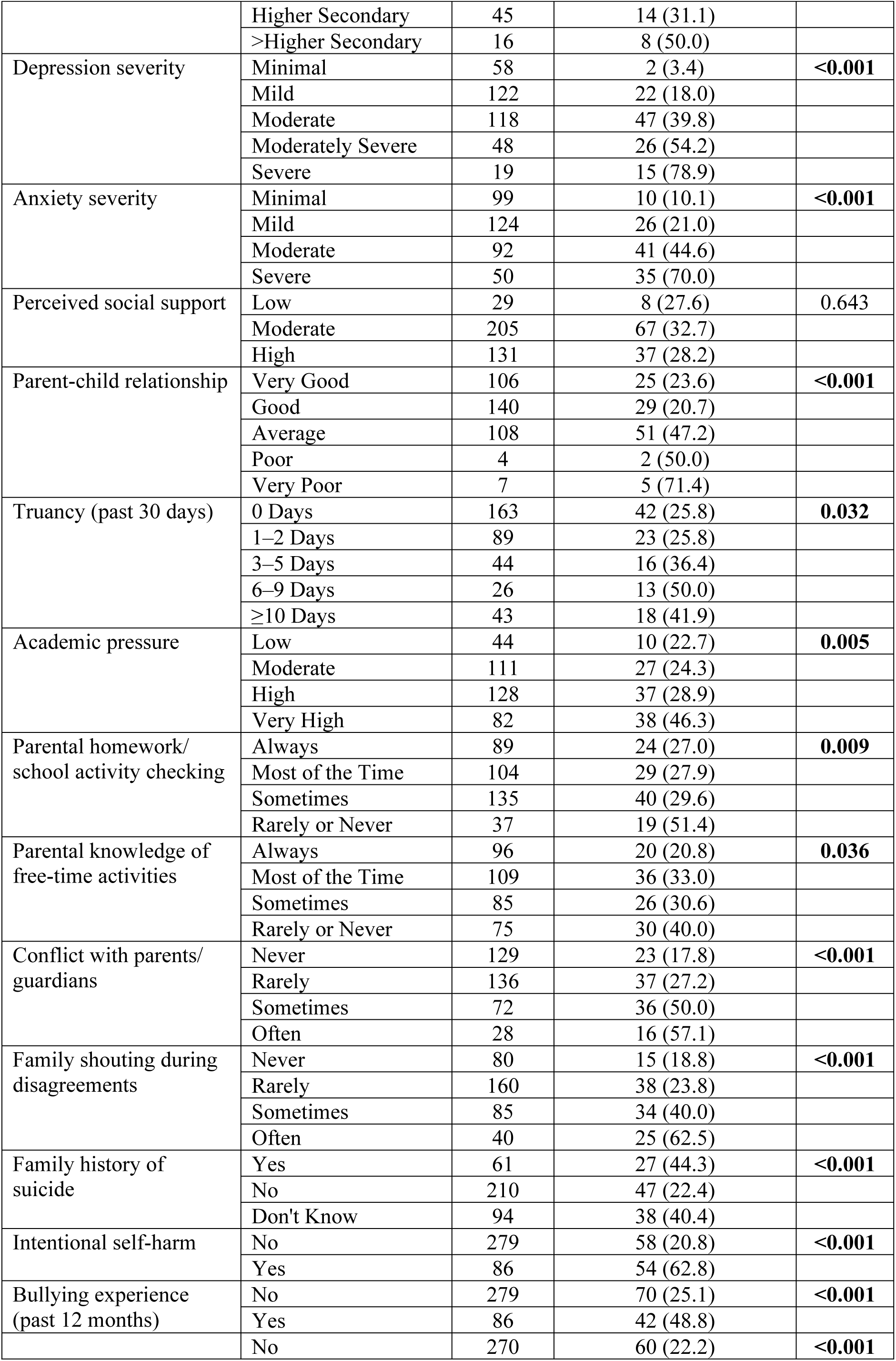
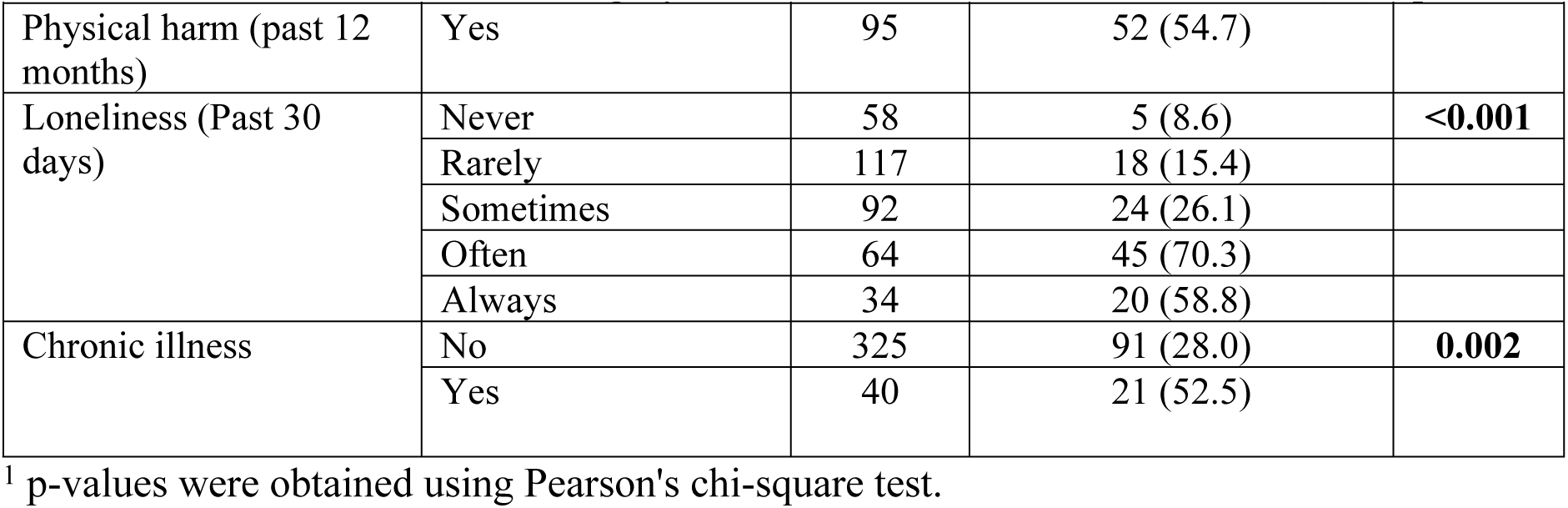
Bivariate associations between participant characteristics and 12-month suicidal ideation among school-going adolescents in Nilphamari District, Bangladesh (N = 365)

### Multivariable associations with suicidal ideation

After adjusting for covariates in the primary multivariable binary logistic regression model, maternal secondary education, depression severity, truancy, intentional self-harm, physical harm, and loneliness were associated with suicidal ideation (Table 3).

**Table 3.** Univariable and multivariable logistic regression analysis of factors associated with 12-month suicidal ideation among school-going adolescents in Nilphamari District, Bangladesh (N = 365)

| Variable | Category | COR (95% CI) | P-value | AOR (95% CI) | P-value |
| --- | --- | --- | --- | --- | --- |
| Residence area | Urban | Reference |  | Reference |  |
|  | Rural | 0.51 (0.29–0.87) | 0.014 | 0.61 (0.24–1.51) | 0.282 |
| Family size | ≤3 | Reference |  | Reference |  |
|  | 4–5 | 0.36 (0.16–0.81) | 0.013 | 0.39 (0.11–1.34) | 0.135 |
|  | ≥6 | 0.33 (0.13–0.81) | 0.015 | 0.26 (0.06–1.07) | 0.062 |
| Mother's education | No Education | Reference |  | Reference |  |
|  | Primary | 0.38 (0.18–0.81) | 0.012 | 0.36 (0.12–1.05) | 0.062 |
|  | Secondary | 0.35 (0.17–0.73) | 0.005 | 0.34 (0.12–0.99) | 0.048 |
|  | Higher Secondary | 0.45 (0.18–1.11) | 0.082 | 0.36 (0.10–1.37) | 0.134 |
|  | >Higher Secondary | 1.00 (0.31–3.22) | 1.000 | 0.56 (0.08–3.99) | 0.566 |
| Depression severity | Minimal | Reference |  | Reference |  |
|  | Mild | 6.16 (1.40–27.17) | 0.016 | 2.30 (0.42–12.63) | 0.340 |
|  | Moderate | 18.54 (4.31–79.64) | <0.001 | 6.05 (1.03–35.38) | 0.046 |
|  | Moderately Severe | 33.09 (7.23–151.36) | <0.001 | 6.86 (1.05–44.62) | 0.044 |
|  | Severe | 105.00 (17.52–629.19) | <0.001 | 7.86 (0.77–80.61) | 0.083 |
| Anxiety severity | Minimal | Reference |  | Reference |  |
|  | Mild | 2.36 (1.08–5.17) | 0.032 | 1.26 (0.42–3.82) | 0.679 |
|  | Moderate | 7.15 (3.31–15.49) | <0.001 | 1.23 (0.38–4.00) | 0.725 |
|  | Severe | 20.77 (8.52–50.60) | <0.001 | 1.64 (0.40–6.80) | 0.492 |
| Parental homework/school activity checking | Always | Reference |  | Reference |  |
|  | Most of the Time | 1.05 (0.56–1.98) | 0.887 | 0.73 (0.28–1.87) | 0.513 |
|  | Sometimes | 1.14 (0.63–2.07) | 0.666 | 1.24 (0.51–3.02) | 0.636 |
|  | Rarely or Never | 2.86 (1.29–6.34) | 0.010 | 1.82 (0.54–6.10) | 0.333 |
| Parental knowledge of free-time activities | Always | Reference |  | Reference |  |
|  | Most of the Time | 1.87 (0.99–3.53) | 0.052 | 0.77 (0.31–1.97) | 0.592 |
|  | Sometimes | 1.67 (0.85–3.29) | 0.134 | 0.89 (0.34–2.34) | 0.815 |
|  | Rarely or Never | 2.53 (1.29–4.98) | 0.007 | 0.80 (0.29–2.24) | 0.670 |
| Parent-child relationship | Very Good | Reference |  | Reference |  |
|  | Good | 0.85 (0.46–1.55) | 0.590 | 0.55 (0.23–1.35) | 0.193 |
|  | Average | 2.90 (1.61–5.21) | <0.001 | 2.13 (0.87–5.22) | 0.097 |
|  | Poor | 3.24 (0.43–24.20) | 0.252 | 0.40 (0.03–4.99) | 0.475 |
|  | Very Poor | 8.10 (1.48–44.34) | 0.016 | 3.77 (0.30–47.20) | 0.304 |
|  | 0 Days | Reference |  | Reference |  |
| Truancy, past 30 days | 1–2 Days | 1.00 (0.56–1.81) | 0.990 | 2.07 (0.82–5.27) | 0.126 |
|  | 3–5 Days | 1.65 (0.81–3.34) | 0.167 | 2.33 (0.80–6.82) | 0.122 |
|  | 6–9 Days | 2.88 (1.24–6.71) | 0.014 | 4.44 (1.24–15.90) | 0.022 |
|  | ≥10 Days | 2.07 (1.03–4.18) | 0.041 | 3.77 (1.34–10.59) | 0.012 |
| Academic pressure | Low | Reference |  | Reference |  |
|  | Moderate | 1.09 (0.48–2.50) | 0.833 | 1.05 (0.30–3.61) | 0.940 |
|  | High | 1.38 (0.62–3.08) | 0.429 | 0.97 (0.29–3.29) | 0.963 |
|  | Very High | 2.94 (1.28–6.72) | 0.011 | 1.48 (0.44–5.01) | 0.525 |
| Conflict with parents/ guardians | Never | Reference |  | Reference |  |
|  | Rarely | 1.72 (0.96–3.10) | 0.070 | 0.84 (0.35–2.02) | 0.699 |
|  | Sometimes | 4.61 (2.42–8.79) | <0.001 | 1.17 (0.43–3.18) | 0.762 |
|  | Often | 6.14 (2.56–14.72) | <0.001 | 1.66 (0.48–5.78) | 0.424 |
| Family shouting during disagreements | Never | Reference |  | Reference |  |
|  | Rarely | 1.35 (0.69–2.64) | 0.380 | 0.85 (0.32–2.26) | 0.750 |
|  | Sometimes | 2.89 (1.42–5.87) | 0.003 | 0.66 (0.22–2.01) | 0.465 |
|  | Often | 7.22 (3.08–16.92) | <0.001 | 0.76 (0.22–2.71) | 0.676 |
| Family history of suicide | Yes | Reference |  | Reference |  |
|  | No | 0.36 (0.20–0.66) | 0.001 | 0.92 (0.36–2.36) | 0.870 |
|  | Don't Know | 0.85 (0.45–1.64) | 0.636 | 1.15 (0.43–3.03) | 0.782 |
| Intentional self-harm | No | Reference |  | Reference |  |
|  | Yes | 6.43 (3.81–10.86) | <0.001 | 2.51 (1.17–5.41) | 0.018 |
| Bullying experience (past 12 months) | No | Reference |  | Reference |  |
|  | Yes | 2.85 (1.73–4.71) | <0.001 | 1.21 (0.55–2.66) | 0.631 |
| Physical harm (past 12 months) | No | Reference |  | Reference |  |
|  | Yes | 4.23 (2.58–6.95) | <0.001 | 2.63 (1.24–5.57) | 0.012 |
| Loneliness (past 30 days) | Never | Reference |  | Reference |  |
|  | Rarely | 1.93 (0.68–5.48) | 0.219 | 1.48 (0.38–5.75) | 0.575 |
|  | Sometimes | 3.74 (1.34–10.46) | 0.012 | 2.10 (0.53–8.32) | 0.291 |
|  | Often | 25.11 (8.68–72.62) | <0.001 | 10.59 (2.53–44.35) | 0.001 |
|  | Always | 15.14 (4.83–47.51) | <0.001 | 2.62 (0.55–12.38) | 0.225 |
| Chronic illness | No | Reference |  | Reference |  |
|  | Yes | 2.84 (1.46–5.53) | 0.002 | 1.66 (0.57–4.86) | 0.354 |

Adolescents whose mothers had attained secondary education had 66% lower odds of suicidal ideation compared to those whose mothers had no formal education (AOR = 0.34, 95% CI: 0.12–0.99; p = 0.048).

Compared to minimal depression, moderate (AOR = 6.05, 95% CI: 1.03–35.38; p = 0.046) and moderately severe depressive symptoms (AOR = 6.86, 95% CI: 1.05–44.62; p = 0.044) were associated with higher odds of suicidal ideation. Feeling lonely “often” was associated with ten-fold higher odds of suicidal ideation (AOR = 10.59, 95% CI: 2.53–44.35; p = 0.001).

Adolescents reporting 6–9 days of truancy during the preceding 30 days had four times higher odds of suicidal ideation than those reporting no truancy (AOR 4.44, 95% CI 1.24–15.90; p=0.022). Those reporting 10 or more days of truancy also had higher odds (AOR 3.77, 95% CI 1.34–10.59; p=0.012).

Both a history of intentional self-harm (AOR = 2.51, 95% CI: 1.17–5.41; p = 0.018) and exposure to physical harm in the past 12 months (AOR = 2.63, 95% CI: 1.24–5.57; p = 0.012) were associated after adjustment with higher odds of suicidal ideation.

After multivariable adjustment, the associations of anxiety, bullying, family-related factors, academic pressure, residence area, family size, and chronic illness were no longer statistically significant (Table 3).

### Sensitivity analysis

A sensitivity analysis using the PHQ-8 score was conducted after excluding PHQ-9 Item 9, which addresses thoughts of death or self-harm and may conceptually overlap with the suicidal ideation outcome. When PHQ-8 was entered as a continuous variable in place of depression severity in the same multivariable logistic regression model, it was not significantly associated with suicidal ideation (AOR = 1.06, 95% CI: 0.96–1.17; p = 0.228). However, the associations for maternal education, truancy, intentional self-harm, physical harm, and loneliness were broadly similar to those observed in the primary model. The full sensitivity analysis results are presented in S2 Table.

### Model fit and discrimination

The primary multivariable model showed adequate calibration (Hosmer-Lemeshow χ^2^ = 11.57, p = 0.172) and good discrimination (AUC = 0.902).

## Discussion

### Principal findings

This study found that 30.7% of school-going adolescents in Nilphamari District reported suicidal ideation during the preceding 12 months, indicating that suicidal ideation was common in this study population. In the primary adjusted analysis, moderate and moderately severe depressive symptoms, higher levels of truancy, intentional self-harm, physical harm, and frequent loneliness were associated with higher odds of suicidal ideation, while secondary maternal education was associated with lower odds. However, depressive symptoms were not statistically significantly associated with suicidal ideation after sensitivity analysis. Although anxiety severity, family-related difficulties, bullying, chronic illness, and several other factors showed significant associations in the bivariate analysis, these associations did not remain statistically significant after adjustment. Taken together, the findings indicate that suicidal ideation among school-going adolescents in this setting was associated with a combination of mental-health difficulties, adverse experiences, and school-related behavioural factors, highlighting the importance of considering these interrelated factors when addressing adolescent suicidal ideation in northern Bangladesh.

### Prevalence of suicidal ideation

This study found a 12-month prevalence of suicidal ideation of 30.7%, indicating a substantial burden among school-going adolescents in northern Bangladesh. This prevalence was higher than estimates reported in several Bangladeshi studies, including 11.7% among school-going adolescents in the national GSHS analysis [9], 19.6% among rural school-going adolescents in southern Bangladesh [11], and 3.2% among adolescents in Sirajganj District [10]. It was also higher than pooled estimates for Bangladesh (15.6%) and South Asia (11.8%) reported in recent systematic reviews [8,17], as well as the pooled estimate of 16.9% among adolescents in low- and middle-income countries [18]. Differences in age composition, study setting, sampling, measurement instruments, and operational definitions of suicidal ideation may partly explain the variation in estimates.

### Depression and anxiety

Depressive symptoms showed an important association with suicidal ideation in the primary analysis. Adolescents with moderate (AOR = 6.05; 95% CI: 1.03–35.38) and moderately severe depressive symptoms (AOR = 6.86; 95% CI: 1.05–44.62) had higher odds of suicidal ideation than those with minimal symptoms. However, this finding was not robust in the sensitivity analysis using the PHQ-8 score after excluding Item 9, which was not significantly associated with suicidal ideation after adjustment. The primary finding should therefore be interpreted cautiously, given the conceptual overlap between the suicidality-related PHQ-9 item and the study outcome. Nevertheless, the observed association in the primary analysis is consistent with previous evidence linking depressive symptoms with suicidal ideation among adolescents [10,11,19–22]. Anxiety severity was also strongly associated with suicidal ideation in the bivariate analysis, with prevalence increasing from 10.1% among adolescents with minimal anxiety to 70.0% among those with severe anxiety (p < 0.001). This pattern is consistent with previous studies reporting significant associations between anxiety symptoms and suicidal ideation among adolescents [9,10,23]. However, the association did not remain statistically significant after adjustment. The attenuation after adjustment may indicate that the observed bivariate association was partly related to overlap with depression and other factors included in the model.

### Behavioural and interpersonal factors

Several behavioural and interpersonal factors were associated after adjustment with suicidal ideation. Adolescents reporting 6–9 days of truancy had more than four-fold higher odds of suicidal ideation (AOR = 4.44; 95% CI: 1.24–15.90), while those reporting 10 or more days had nearly four-fold higher odds (AOR = 3.77; 95% CI: 1.34–10.59). A history of intentional self-harm was also associated with higher odds of suicidal ideation (AOR = 2.51; 95% CI: 1.17–5.41), as was experiencing physical harm during the preceding 12 months (AOR = 2.63; 95% CI: 1.24–5.57). These findings are consistent with previous evidence linking school absenteeism, self-harm, and exposure to violence with suicidal ideation among adolescents [24–29]. Loneliness showed the strongest association, with adolescents who often experienced loneliness having more than ten-fold higher odds of suicidal ideation than those who never felt lonely (AOR = 10.59; 95% CI: 2.53–44.35). This finding is consistent with evidence identifying loneliness as an important factor associated with suicidal ideation among adolescents [9,30–35]. Together, these findings highlight the importance of addressing school disengagement, self-harm, exposure to physical harm, and social isolation in adolescent suicide-prevention efforts.

### Family and educational factors

Maternal education was the only socio-demographic factor that remained statistically significantly associated with suicidal ideation after adjustment. Adolescents whose mothers had attained secondary education had 66% lower odds of suicidal ideation compared to those whose mothers had no formal education (AOR = 0.34, 95% CI: 0.12–0.99; p = 0.048). However, the absence of a consistent gradient across maternal education categories warrants cautious interpretation. This finding is consistent with evidence suggesting that higher parental education may be associated with better parent–child communication and more supportive parenting practices, which may in turn contribute to a lower likelihood of suicidal ideation among adolescents. [14,16].

Several family and interpersonal factors including parent–child relationship, conflict with parents or guardians, family shouting during disagreements, and family history of suicide were significantly associated with suicidal ideation in the bivariate analysis but did not remain statistically significant after adjustment. The attenuation of these associations may reflect overlap with other psychosocial and behavioural factors included in the multivariable model, particularly depression, self-harm, physical harm, truancy, and loneliness. This suggests that the apparent influence of family and interpersonal factors may be partly accounted for by co-occurring psychological and behavioural vulnerabilities.

### Implications

The findings highlight the need for early identification of adolescents experiencing mental-health and psychosocial difficulties, particularly through school-based screening and support. Schools should pay particular attention to adolescents with self-harm, frequent loneliness, repeated truancy, or exposure to physical harm, while strengthening referral pathways for appropriate mental-health care. Engaging families and strengthening supportive parent–adolescent relationships may further contribute to suicide-prevention efforts. These approaches may be particularly relevant for strengthening adolescent mental-health and suicide-prevention services in northern Bangladesh.

### Limitations

This study has several limitations. First, because the study was cross-sectional, it was not possible to determine the temporal relationship between suicidal ideation and the factors examined. Second, the study was conducted in only two upazilas of Nilphamari District and included selected co-educational schools and colleges; therefore, the findings may not be generalisable to all school-going adolescents in Bangladesh, particularly those attending single-sex institutions or living in other regions. Third, participants were recruited from students present on the scheduled data-collection days, which may have introduced selection bias related to school absence. Finally, suicidal ideation and other sensitive experiences were self-reported, so some responses may have been affected by recall or social-desirability bias. Although a range of relevant factors was included in the analysis, some other factors were not assessed and may have influenced the observed associations.

## Conclusion

Suicidal ideation was common among school-going adolescents in northern Bangladesh. Truancy, intentional self-harm, physical harm and frequent loneliness were associated after adjustment with suicidal ideation, while secondary maternal education was associated with lower odds. Although moderate and moderately severe depressive symptoms were associated with suicidal ideation in the primary analysis, this finding was not robust in the PHQ-8 sensitivity analysis. These findings underscore the need for early, school-based identification and support for adolescents experiencing mental-health and psychosocial difficulties, alongside family engagement and strengthened suicide-prevention services in northern Bangladesh.

## Data Availability

The de-identified dataset underlying the findings reported in this study is publicly available in Zenodo at https://zenodo.org/records/22981953

## Acknowledgements

The authors are grateful to all the adolescent participants who voluntarily took part in this study and shared their experiences through the survey. We also express gratitude to the teachers and staff of the participating institutions.

## Supporting information

S1 Checklist. STROBE statement – checklist of items that should be included in reports of 453 cross-sectional studies.

S1 Table. Additional bivariate associations between selected participant characteristics and 12-month suicidal ideation among school-going adolescents in Nilphamari District, Bangladesh (N = 365)

S2 Table. Sensitivity analysis using the PHQ-8 score in the multivariable logistic regression model for 12-month suicidal ideation

